# Risk of incident cardiovascular disease among patients with gastrointestinal disorder: prospective cohort study of 340,862 individuals

**DOI:** 10.1101/2023.04.09.23288329

**Authors:** Jie Chen, Yuhao Sun, Tian Fu, Shiyuan Lu, Wenming Shi, Jianhui Zhao, Sen Li, Xue Li, Shuai Yuan, Susanna C. Larsson

## Abstract

**Background and Aims:** The associations between gastrointestinal diseases (GIs) and cardiovascular disease (CVD) were unclear. We conducted a prospective cohort study to explore their associations.

**Methods:** This study included 340,862 individuals without baseline CVD from the UK Biobank cohort. Individuals with and without GIs were followed up until the ascertainment of incident CVDs, including coronary heart disease (CHD), cerebrovascular disease (CeVD), and peripheral artery disease (PAD). The diagnosis of diseases was confirmed with combination of the nationwide inpatient data, primary care data, and cancer registries. A multivariable Cox proportional hazard regression model was used to estimate the associations between GIs and the risk of incident CVD.

**Results:** During a median follow-up of 12.4 years, 28,787 incident CVD cases were diagnosed. Individuals with GIs had an elevated risk of CVD (hazard ratio 1.38; 95% confidence interval 1.35-1.42, *P*<0.001). Eleven out of fifteen GIs were associated with an increased risk of CVD after Bonferroni-correction, including cirrhosis, non-alcoholic fatty liver disease, gastritis and duodenitis, irritable bowel syndrome, gastroesophageal reflux disease, peptic ulcer, celiac disease, pancreatitis, diverticular disease, biliary disease, and appendicitis. The associations were stronger among women, individuals aged ≤ 60 years, and those with body mass index ≥ 25 kg/m^2^.

**Conclusions:** This large-scale prospective cohort study revealed the associations of GIs with an increased risk of incident CVD, in particular CHD and PAD. These findings support the reinforced secondary CVD prevention among patients with gastrointestinal disorders.

## Introduction

A gut-heart connection has been proposed to explain an excessive risk of cardiovascular disease (CVD) among individuals with gastrointestinal disorders.^1^ This axis has been supported by several pathways, such as blood pressure changes via glucagon-like peptide-1 receptor,^2^ alterations in gut microbiome and related metabolites,^3^ and increased oxidative stress and inflammation^4^. Furthermore, common gastrointestinal diseases,^5^ including reflux esophagitis,^6^ non-alcoholic fatty liver disease,^7^ gallstone disease,^8^ and some bowel diseases,^9-12^ were found to be positively associated with risk of CVD in several population-based epidemiological studies. However, these associations were not found in other studies,^13-16^ and there are limited data on other gastrointestinal diseases in relation to the risk of CVD. Thus, the associations between gastrointestinal diseases and the development of CVD remain unclear. In addition, sex difference was observed in the associations between certain gastrointestinal diseases and the risk of subsequent CVD with a particularly greater risk in women.^11^ Whether this sex specific finding can be generalized to the associations between different gastrointestinal disorders and CVD risk has not been assessed in a comprehensive manner.

A clear appraisal of the associations between gastrointestinal disorders and incident CVD risk can not only deepen the etiological understanding of the development of CVD but also lend evidence support for the secondary prevention of CVD in patients with gastrointestinal diseases. Therefore, we conducted this cohort study to examine the associations of a broad range of gastrointestinal diseases with the risk of incident CVD. We further differentiated these associations into subgroups defined by age, sex, and body mass index (BMI) with the aim of identifying the vulnerable group for a better prevention purpose.

## Methods

### Study population

The analysis was based on data from the UK Biobank study, an ongoing cohort study that recruited over 500,000 individuals aged 40-69 years old from 22 centres in the UK between 2006 and 2010. In UK Biobank, phenotypic information is derived from self-administrative questionnaires, computer-assisted interviews, physical checks, and blood, urine, and saliva samples. Health outcome data are available via a linkage to national datasets. We removed 158,586 individuals with baseline cardiovascular disease and 2629 newly onset cases within 1 year of follow-up, resulting in 340,862 participants included in the analysis. The UK Biobank study had been approved by the North West– Haydock Research Ethics Committee (REC reference: 21/NW/0157). All participants had signed an electronic consent. The study was conducted by the STROBE checklist.

### Definitions of Gastrointestinal Diseases

The study included fifteen gastrointestinal endpoints that were defined by International Classification of Disease (ICD) codes (**Table S1**) with data from the nationwide inpatient dataset, primary care dataset, and cancer registries. Fifteen gastrointestinal diseases included oesophageal diseases (gastroesophageal reflux disease and Barrett’s oesophagus), gastric and bowel diseases (gastritis and duodenitis, celiac disease, peptic ulcer, Crohn’s disease, ulcerative colitis, irritable bowel syndrome, and intestinal diverticular disease), pancreatitis (acute and chronic), biliary disease (cholangitis, cholecystitis, and cholelithiasis), liver diseases (non-alcoholic fatty liver disease and chronic liver cirrhosis), appendicitis, and overall gastrointestinal cancer (oesophageal, gastric, small intestinal, colorectal, pancreatic, gallbladder, and liver cancers). The accuracy of ICD codes has been proven to be satisfying in the annual report of the Audit Commission for Local Authorities of the National Health Service in England and Wales.^17^

### Ascertainment of CVDs and follow-up

We also used ICD codes to define incident overall CVDs (including coronary heart disease [CHD], cerebrovascular disease [CeVD], and peripheral artery disease [PAD]) (**Table S2**) with diagnostic data from nationwide inpatient dataset and primary care dataset. For baseline CVDs, we additionally used self-report diagnosis (**Table S2**) reviewed by nurses to remove individuals with baseline CVD in a conservative way. Participants were followed up from recruitment date to the date of first diagnosis, death, loss to follow-up, the last date of hospital admission, or end of the follow-up whichever came first.

### Assessment of Covariates

We treated age at recruitment, sex, Townsend deprivation index, BMI, education attainment, ethnicity, smoking status, alcohol consumption status, physical activity, and adherence to a healthy diet as important covariates. Except for BMI (weight/height^2^) with data measured by trained nurses at clinical visits, information on other covariates was obtained from the baseline touch-screen questionnaires. A healthy diet (a cardioprotective diet) was constructed by intakes of seven food groups (fruits, vegetables, fish, whole grains, refined grains, processed meats, and unprocessed red meats) with data from food frequency questionnaires.^18^ We additionally extracted data on baseline hypertension, hyperlipemia, diabetes, metabolic syndrome, family history of CVDs, INFLA (immune biomarkers and an aggregated inflammation)-score,^19^ systemic immune-inflammation index,^20^ systemic inflammation response index^21^, use of proton pump inhibitors, and depression symptoms. Detailed definitions and data sources for all the aforementioned covariates are shown in **Table S3.**

### Statistical Analysis

The missing rate of covariates ranged from 0.1% for Townsend deprivation index to 9.3% for INFLA-score (**Table S4**). We imputed continuous variables with sex-specific median and categorical variables with plural for those with less than 3% missing and with the indicator for those with more than 3% missing. Cox proportional hazards regression model, treating age as a timescale, was used to estimate the associations between 15 gastrointestinal diseases and the risk of incident CVDs with adjustment for sex, Townsend deprivation index, BMI, education attainment, ethnicity, smoking status, alcohol consumption, physical activity, and adherence to a healthy diet. We also examined the associations by major locations of gastrointestinal diseases. We tested the multiplicative interactions of gastrointestinal diseases with age (≤60 or >60 years old), sex (women and men), and BMI (≥25 or <25 kg/m^2^) and stratified the associations by these factors. Several sensitivity analyses were performed to test the robustness of the findings: 1) additionally adjusting for INFLA-score, systemic immune-inflammation index, systemic inflammation response index, baseline hypertension, hyperlipemia, and diabetes, metabolic syndrome, family history of CVDs, use of proton pump inhibitors, and depression symptoms; 2) excluding incident CVDs within 2 years follow-up; and 3) excluding incident cases with gastrointestinal diseases; 4) for GI cancers, adding a competing risk model with death as a competing event. We used Bonferroni correction to adjust for multiple testing. The association with a *P-*value <0.003 (0.05/16 exposures) was deemed significant. The association with a Bonferroni-adjusted *P-*value >0.05 and a nominal *P-*value <0.05 was regarded as suggestive. All tests were two-sided and performed using R software, version 4.2.1.

## Results

During a median follow-up of 12.4 years, 28,787 incident overall CVD cases (including 20,487 CHD cases, 8326 CeVD cases, and 2206 PAD cases) were diagnosed. **Table 1** shows the baseline characteristics by incident CVD status. Compared to individuals without incident CVD, those with incident CVD had an unhealthier lifestyle, and more baseline comorbidities and gastrointestinal diseases.

**Table 1.** Baseline characteristics of study participants by incident status of cardiovascular diseases

| **Characteristic** | **Overall** | **Non-CVD** | **Incident CVD** |
| --- | --- | --- | --- |
| N | 340862 | 312075 | 28787 |
| Age (mean (SD)) | 55.10 (8.14) | 54.75 (8.11) | 58.91 (7.45) |
| Women (%) | 197396 (57.9) | 184895 (59.2) | 12501 (43.4) |
| TDI (mean (SD)) | -1.41 (3.03) | -1.43 (3.02) | -1.21 (3.13) |
| BMI (mean (SD)) | 26.63 (4.38) | 26.56 (4.36) | 27.39 (4.48) |
| College degree and above (%) | 119937 (35.2) | 112027 (35.9) | 7910 (27.5) |
| White (%) | 322066 (94.5) | 294773 (94.5) | 27293 (94.8) |
| Ever smoking (%) | 144940 (42.5) | 130071 (41.7) | 14869 (51.7) |
| Current drinker (%) | 316101 (92.7) | 289859 (92.9) | 26242 (91.2) |
| Regular physical activity (%) | 273986 (80.4) | 251304 (80.5) | 22682 (78.8) |
| Adherence to a healthy diet (%) | 237281 (69.6) | 218484 (70.0) | 18797 (65.3) |
| Metabolic syndrome (%) | 24618 (7.2) | 21685 (6.9) | 2933 (10.2) |
| Hypertension, hyperlipidaemia, or diabetes (%) | 35858 (10.5) | 30700 (9.8) | 5158 (17.9) |
| Family of CVD (%) | 171146 (50.2) | 154863 (49.6) | 16283 (56.6) |
| Use of PPI (%) | 23606 (6.9) | 20223 (6.5) | 3383 (11.8) |
| Depression symptoms (%) | 17132 (5.0) | 15379 (4.9) | 1753 (6.1) |
| Inflammatory indexes |  |  |  |
| INFLA (mean (SD)) | -0.54 (5.75) | -0.62 (5.75) | 0.29 (5.75) |
| SII (mean (SD)) | 585.00 (333.10) | 583.82 (329.81) | 597.79 (366.71) |
| SIRI (mean (SD)) | 1.05 (0.97) | 1.05 (0.98) | 1.15 (0.82) |
| **Gastrointestinal diseases** |  |  |  |
| Barrett's oesophagus (%) | 1007 (0.3) | 871 (0.3) | 136 (0.5) |
| GERD (%) | 21906 (6.4) | 19007 (6.1) | 2899 (10.1) |
| Gastritis and duodenitis (%) | 13377 (3.9) | 11504 (3.7) | 1873 (6.5) |
| Celiac disease (%) | 1737 (0.5) | 1556 (0.5) | 181 (0.6) |
| Peptic ulcer (%) | 6907 (2.0) | 5861 (1.9) | 1046 (3.6) |
| UC (%) | 1543 (0.5) | 1386 (0.4) | 157 (0.5) |
| CD (%) | 2853 (0.8) | 2556 (0.8) | 297 (1.0) |
| IBS (%) | 16110 (4.7) | 14437 (4.6) | 1673 (5.8) |
| Diverticulum (%) | 8362 (2.5) | 7172 (2.3) | 1190 (4.1) |
| Pancreatitis (%) | 1125 (0.3) | 971 (0.3) | 154 (0.5) |
| NAFLD (%) | 573 (0.2) | 476 (0.2) | 97 (0.3) |
| Cirrhosis (%) | 848 (0.2) | 721 (0.2) | 127 (0.4) |
| Biliary disease (%) | 10289 (3.0) | 9173 (2.9) | 1116 (3.9) |
| Appendicitis (%) | 5101 (1.5) | 4605 (1.5) | 496 (1.7) |
| Gastrointestinal cancer (%) | 2206 (0.6) | 1919 (0.6) | 287 (1.0) |
| Any gastrointestinal disease (%) | 69388 (20.4) | 61311 (19.6) | 8077 (28.1) |

Abbreviations: BMI, body mass index; CD, Crohn's disease; CeVD, cerebrovascular disease; CHD, coronary heart disease; CI, confidence interval; CVD, cardiovascular disease; GERD, gastroesophageal reflux disease; HR-hazard ratio; IBS, irritable bowel syndrome; NAFLD, non-alcoholic fatty liver disease; PAD, peripheral artery disease; PPI, proton pump inhibitors; SD, standard deviation; TDI, Townsend deprivation index; UC, ulcerative colitis.

### Overall CVD

Having any gastrointestinal disease was associated with an elevated risk of overall CVD (hazard ratio [HR] 1.38; 95% confidence interval [CI] 1.35-1.42, *P*<0.001). Eleven out of fifteen gastrointestinal diseases were associated with an increased risk of overall CVD after multiple testing corrections; the HR ranged from 1.19 for biliary disease and for appendicitis to 1.65 and 1.74 for the liver diseases non-alcoholic fatty liver disease and cirrhosis, respectively (**Figure 1**). There were suggestive associations (*P*<0.02) of three gastrointestinal diseases (i.e., ulcerative colitis, gastrointestinal cancer, and Crohn’s disease) and no association of Barrett’s oesophagus with overall CVD risk. Gastrointestinal diseases by location were associated with a high risk of incident CVD (**Table 2**). The HR of CVD ranged from 1.19 (95% CI 1.08-1.30, *P*<0.001) for diseases of appendix to 1.40 (95% CI 1.27-1.53, *P*<0.001) for liver diseases.

**Figure 1.**
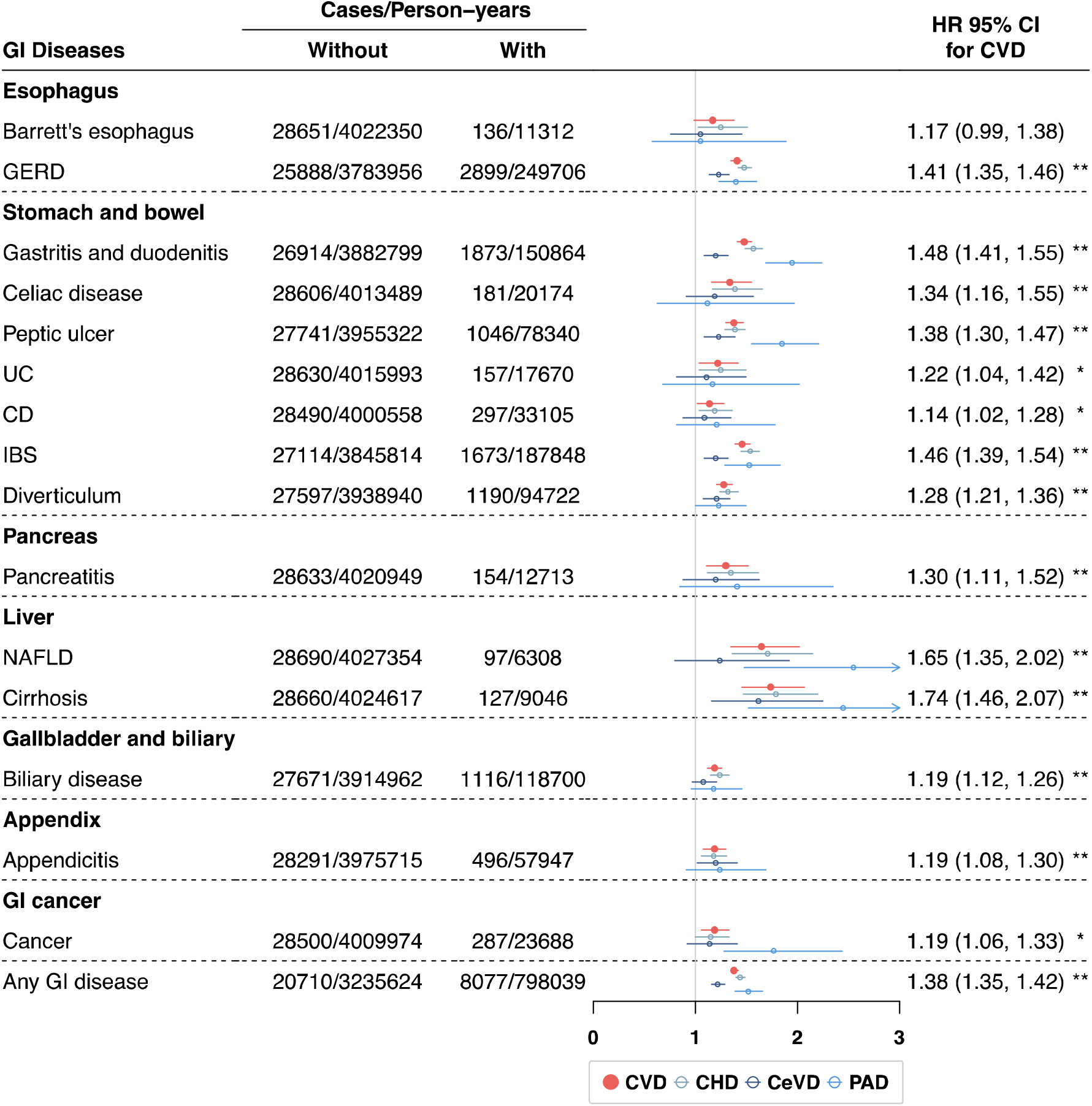
Hazard ratios for the associations of gastrointestinal diseases with incident cardiovascular disease. The associations were adjusted for age, sex, Townsend deprivation index, body mass index, education, ethnicity, smoking status, drinking status, physical activity, and adherence to a healthy diet. * *P*<0.05 and ** *P*<0.05/16 after Bonferroni correction. Abbreviations: CD, Crohn’s disease; CeVD, cerebrovascular disease; CHD, coronary heart disease; CI, confidence interval; GERD, gastroesophageal reflux disease; CVD, cardiovascular disease; GI, gastrointestinal disease; HR-hazard ratio; IBS, irritable bowel syndrome; NAFLD, non-alcoholic fatty liver disease; PAD, peripheral artery disease; UC, ulcerative colitis.

**Table 2.** Hazard ratios of incident cardiovascular disease for different locations of gastrointestinal diseases

| Disease location | Cases/Person-years |  | HR (95% CI) | P-value |
| --- | --- | --- | --- | --- |
|  | Without | With |  |  |
| Oesophageal | 25819/3777706 | 2968/255956 | 1.40 (1.35, 1.45) | <0.001 |
| Stomach and bowel | 23465/3526832 | 5322/506830 | 1.39 (1.35, 1.43) | <0.001 |
| Pancreas | 28628/4020255 | 159/13407 | 1.28 (1.09, 1.49) | 0.002 |
| Liver | 28331/3999260 | 456/34402 | 1.40 (1.27, 1.53) | <0.001 |
| Biliary | 27666/3914646 | 1121/119016 | 1.19 (1.12, 1.26) | <0.001 |
| Appendix | 28291/3975715 | 496/57947 | 1.19 (1.08, 1.30) | <0.001 |

The associations were adjusted for age, sex, Townsend deprivation index, body mass index, education, ethnicity, smoking status, drinking status, physical activity, and adherence to a healthy diet. Abbreviations: HR-hazard ratio, CI-confidence interval.

There were multiplicative interactions observed for some gastrointestinal diseases with age, sex, or BMI (**Figure 2 and Table S6**). In stratified analysis, the associations between any gastrointestinal disease and incident CVD appeared to be stronger among women (women: HR 1.48; 95% CI 1.43-1.54, *P*<0.001; man: HR 1.30; 95% CI 1.25-1.35, *P* <0.001), those with baseline age ≤ 60 years (≤ 60: HR 1.46; 95% CI 1.40-1.51, *P* <0.001; > 60: HR 1.32; 95% CI 1.27-1.36, *P* <0.001), and those with BMI ≥ 25 kg/m^2^ (≥25: HR 1.46; 95% CI 1.39-1.53, *P* <0.001; < 25: HR 1.36; 95% CI 1.32-1.41, *P* <0.001).

**Figure 2.**
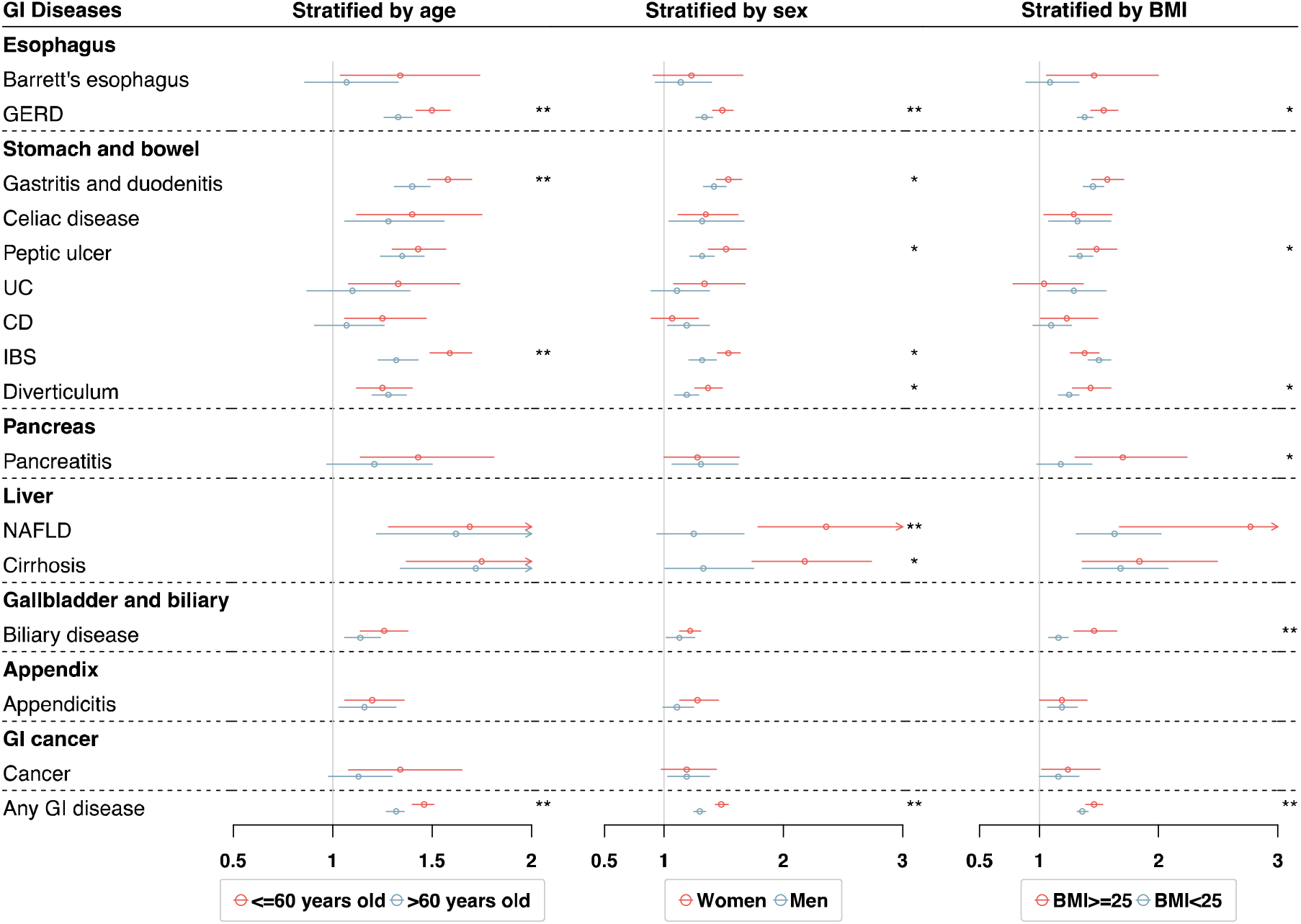
Hazard ratios for the associations of gastrointestinal diseases with incident cardiovascular disease, stratified by age, sex, and body mass index (BMI). The associations were adjusted for age, sex, Townsend deprivation index, body mass index, education, ethnicity, smoking status, drinking status, physical activity, and adherence to a healthy diet. * *P* for interaction<0.05 and ** *P* for interaction<0.05/16 after Bonferroni correction. Abbreviations: BMI, body mass index; CD, Crohn’s disease; CI, confidence interval; GERD, gastroesophageal reflux disease; GI, gastrointestinal disease; IBS, irritable bowel syndrome; NAFLD, non-alcoholic fatty liver disease; UC, ulcerative colitis.

The associations were stronger among individuals with a baseline age ≤ 60 years for gastroesophageal reflux disease, gastritis and duodenitis, and irritable bowel syndrome in relation to incident CVD risk. The associations were stronger in women for gastroesophageal reflux disease, gastritis and duodenitis, peptic ulcer, irritable bowel syndrome, diverticulum, non-alcoholic fatty liver disease, and cirrhosis in relation to CVD. Likewise, the associations of gastroesophageal reflux disease, peptic ulcer, diverticular disease, pancreatitis, and biliary disease with CVD risk were more pronounced in participants with BMI ≥ 25 kg/m^2^.

### CHD, CeVD, and PAD

All studied gastrointestinal diseases were significantly or suggestively associated with an elevated risk of CHD (**Table 3**). The HR of CHD ranged from 1.15 (95% CI 1.00, 1.33, *P*=0.047) for gastrointestinal cancer to 1.79 (95% CI 1.47, 2.20, *P*<0.001) for cirrhosis. Five and seven gastrointestinal diseases were associated with an increased risk of CeVD and PAD, respectively, after adjusting for multiple testing (**Table 3**). There were two suggestive associations of cirrhosis and appendicitis with the risk of CeVD (**Table 3**). The associations for CeVD were overall moderate, except for the association between cirrhosis and CeVD (HR 1.62; 95% CI 1.16-2.25, *P*=0.005). Compared to that of CHD, the associations for PAD were generally larger with the HR of 2.55 (95% CI 1.48-4.40, *P*<0.001) and 2.45 (95% CI 1.52-3.96, *P*=0.001) specifically for the associations of cirrhosis and non-alcoholic fatty liver disease, respectively. The associations remained consistent for the subtypes of CHD (i.e., myocardial infarction) and CeVD (i.e., stroke and transient ischemic attack) with larger CIs (**Table S5**).

**Table 3.** Hazard ratios for the associations of gastrointestinal diseases with incident coronary heart disease (CHD), cerebrovascular disease (CeVD), and peripheral artery disease (PAD)

| Gastrointestinal diseases | CHD |  | CeVD |  | PAD |  |
| --- | --- | --- | --- | --- | --- | --- |
|  | HR (95% CI) | P-value | HR (95% CI) | P-value | HR (95% CI) | P-value |
| **Oesophagus** |  |  |  |  |  |  |
| Barrett's oesophagus | 1.25 (1.03, 1.51) | 0.025 | 1.05 (0.76, 1.46) | 0.767 | 1.05 (0.58, 1.89) | 0.884 |
| GERD | 1.48 (1.42, 1.55) | <0.001 | 1.23 (1.14, 1.33) | <0.001 | 1.40 (1.23, 1.60) | <0.001 |
| **Stomach and bowel** |  |  |  |  |  |  |
| Gastritis and duodenitis | 1.57 (1.49, 1.66) | <0.001 | 1.20 (1.09, 1.32) | <0.001 | 1.95 (1.69, 2.24) | <0.001 |
| Celiac disease | 1.39 (1.17, 1.66) | <0.001 | 1.19 (0.91, 1.57) | 0.202 | 1.12 (0.63, 1.97) | 0.704 |
| Peptic ulcer | 1.39 (1.29, 1.49) | <0.001 | 1.23 (1.09, 1.39) | 0.001 | 1.85 (1.55, 2.21) | <0.001 |
| UC | 1.25 (1.04, 1.50) | 0.02 | 1.11 (0.82, 1.50) | 0.501 | 1.17 (0.68, 2.02) | 0.568 |
| CD | 1.19 (1.04, 1.36) | 0.012 | 1.09 (0.88, 1.35) | 0.432 | 1.21 (0.82, 1.78) | 0.335 |
| IBS | 1.54 (1.45, 1.63) | <0.001 | 1.20 (1.09, 1.32) | <0.001 | 1.53 (1.29, 1.83) | <0.001 |
| Diverticulum | 1.32 (1.24, 1.42) | <0.001 | 1.21 (1.08, 1.34) | 0.001 | 1.23 (1.00, 1.50) | 0.045 |
| **Pancreas** |  |  |  |  |  |  |
| Pancreatitis | 1.35 (1.12, 1.62) | 0.002 | 1.20 (0.88, 1.63) | 0.247 | 1.41 (0.85, 2.35) | 0.182 |
| **Liver** |  |  |  |  |  |  |
| NAFLD | 1.71 (1.36, 2.15) | <0.001 | 1.24 (0.80, 1.92) | 0.343 | 2.55 (1.48, 4.40) | 0.001 |
| Cirrhosis | 1.79 (1.47, 2.20) | <0.001 | 1.62 (1.16, 2.25) | 0.005 | 2.45 (1.52, 3.96) | <0.001 |
| **Gallbladder and biliary** |  |  |  |  |  |  |
| Biliary disease | 1.24 (1.15, 1.33) | <0.001 | 1.08 (0.97, 1.21) | 0.176 | 1.18 (0.96, 1.46) | 0.120 |
| **Appendix** |  |  |  |  |  |  |
| Appendicitis | 1.18 (1.06, 1.31) | 0.002 | 1.20 (1.02, 1.41) | 0.032 | 1.24 (0.91, 1.69) | 0.179 |
| **Gastrointestinal cancer** |  |  |  |  |  |  |
| Cancer | 1.15 (1.00, 1.33) | 0.047 | 1.14 (0.92, 1.41) | 0.248 | 1.77 (1.28, 2.44) | <0.001 |
| **Any above disease** | 1.44 (1.40, 1.49) | <0.001 | 1.22 (1.16, 1.29) | <0.001 | 1.52 (1.39, 1.66) | <0.001 |

### Sensitivity analyses

The observed associations between gastrointestinal diseases and incident overall CVD risk remained stable in the sensitivity analysis additionally adjusted for different inflammatory indexes (**Table S7**) and in the sensitivity analysis additionally adjusted for common comorbidities, metabolic syndrome, family history of CVD, and depression symptoms (**Table S8**). Although the association remains significant, the effect value is attenuated after adjusting for PPI (**Table S8**). The associations remained with slight attenuation in the analysis excluding incident CVD diagnosed within a 2-year follow-up (**Table S8**) and excluding incident gastrointestinal disease cases (**Table S9**). After including death as a competing risk event, GI cancer was associated with a higher risk of CVD (HR 1.45; 95% CI 1.29-1.63, *P*<0.001), CHD (HR 1.37; 95% CI 1.19-1.58, *P*<0.001), CeVD (HR 1.46; 95% CI 1.18-1.82, *P*=0.001), and PAD (HR 2.27; 95% CI 1.65-3.13, *P*<0.001).

## Discussion

This prospective cohort study of 340,862 participants found associations between a wide range of gastrointestinal diseases and increased risk of incident CVD, with the most pronounced associations for liver diseases, including non-alcoholic fatty liver disease and cirrhosis. The associations were robust in a series of sensitivity analyses adjusting for different inflammatory indexes. For major subtypes of CVD, the associations were overall consistent for CHD and PAD but weaker for CeVD. There were interaction effects of certain gastrointestinal diseases, like gastroesophageal reflux disease, with age, sex, and BMI on the risk of CVD. The associations appeared to be stronger in women, individuals with baseline age ≤ 60 years, and overweight and obese participants.

Among all studied gastrointestinal diseases, this study observed most pronounced associations between liver diseases and a higher risk of CVD. The observed associations are in line with most ^7,22,23^ but not all previous findings^16^. In a large-scale Danish cohort of the general population, both increased liver fat content and clinically diagnosed non-alcoholic fatty liver disease were associated with a higher risk of ischemic heart disease.^24^ However, the observational association for liver fat content was not confirmed in Mendelian randomization (MR) analysis where liver fat was proxied by genetic variants in three protein encoding genes.^24^ In recent MR studies, the positive associations were identified between non-alcoholic fatty liver disease and the risk of CHD and myocardial infarction using genetic instruments for non-alcoholic fatty liver disease defined by increased alanine aminotransferase levels, imaging, or biopsy information.^25,26^ All these findings may imply liver disease as a causal risk factor for the development of CVD, in particular CHD. This link may be mediated by multiple traditional cardiovascular risk factors, such as obesity, diabetes, metabolic syndrome, hyperlipidaemia, and hypertension.^27^ In our study, the associations between liver diseases and CVD risk remained after adjusting for above factors, which indicates more alternative pathways as potential mediators. Of note, the association between non-alcoholic fatty liver disease and CVD risk was also observed in paediatric and young adults (HR=5.27, 95% CI 1.96-14.19) with a much larger strength compared to that among middle-aged adults (HR=1.63, 95% CI 1.56-1.70).^28^ Even the association was less precise due to a smaller sample size in paediatric and young adults, this excessive risk of CVD among paediatric and young adult non-alcoholic fatty liver disease may be caused by a longer exposure, which alarms the significance of early secondary prevention for CVD in this high-risk population.

Our findings on some oesophageal and bowel diseases in relation to CVD risk are overall in line with previous studies that identified positive associations of reflux esophagitis,^6,29^ peptic ulcer,^30^ celiac disease,^12,31,32^, and diverticular disease,^9,33^ with CVD risk. However, a population-based cohort study suggested that gastroesophageal reflux disease might not be an independent risk factor for myocardial infarction since the risk of myocardial infarction in patients with gastro-oesophageal reflux disease merely increased in the immediate days after diagnosis. In our study removing all baseline CVD patients including those with myocardial infarction, the association persisted in the analysis excluding incident CVD cases within 2-year follow-up. In addition, the causality of the associations between gastroesophageal reflux disease and several CVDs was strengthened in an MR study.^34^ This study also identified positive associations of gastritis and duodenitis and irritable bowel syndrome with the risk of CVD, which were explored in few studies with conflicting findings.^15,35^ Future studies are warranted to confirm these associations. There was a moderate positive association between Barrett’s oesophagus and CVD risk,^36^ which was not clearly identified in this study. The possible reason of this discrepancy may be an inadequate power of the current analysis due to a small number of baseline Barrett’s oesophagus patients.

Diabetes as a major complication of pancreatitis have been associated with an increased risk of CVD,^37^ which supports the link from pancreatitis to an excessive risk of CVD. This link was also directly identified in some previous cohort studies^38,39^ as well as in our findings. Gallstone disease has been consistently associated with an increased risk of CVD.^8,40,41^ For other biliary diseases, limited data have been collected. Similarly, a few studies have been conducted to explore the association between appendicitis and the risk of CVD. One prospective matched cohort study in a Swedish population found that appendectomy before age 20 was associated with a high risk of acute myocardial infarction.^42^ As for gastrointestinal cancers in relation to CVD risk, studies have been focused on colorectal cancer and less on other cancer of gastroenterological system.^43^ Patients with colorectal cancer may have around 3 times risk of developing CVD compared to individuals without the history of colorectal cancer.^43^ This study revealed a suggestively moderate association between any gastrointestinal cancer and the risk of CVD. More studies are needed to verify our findings.

Interactions of gastrointestinal diseases with age, sex, and BMI on the risk of CVD were scarcely investigated. A systematic review and meta-analysis of nine studies found that women had a higher risk of cardiovascular morbidity by exposure to inflammatory bowel disease compared to men.^11^ Even though this sex-specific association was not found for inflammatory bowel disease in our study, our analysis identified interactions of several other gastrointestinal diseases with age, sex, or BMI. These findings are of great clinical and public health implications for selecting the high-risk population for secondary prevention.

Several underlying pathways have been proposed to explain the positive association between gastrointestinal diseases and CVD risk. First, the changes in metabolic features among patients with gastrointestinal disorders may increase the risk of developing CVD.^27^ Our analysis adjusted for most metabolic surrogates and still found many positive associations, which indicates that there are other major alternative underlying pathways explained these positive links. Second, increased oxidative stress and inflammation in gastrointestinal disease, in particular liver impairment, may facilitate the development of CVD.^4^ Our analysis partly supported this hypothesis by observing slightly attenuated associations after additionally adjusting for different inflammatory indexes, which are based on the levels of C-reactive protein and white cells counts.^19-21^ Other inflammatory pathways may also mediate these associations, such as interleukin-18^44^ and tumour necrosis factor^45^. Third, alternations of gut microbiota and their metabolites in patients with gastrointestinal diseases may explain the excessive risk of CVD, ^3,46^ particularly short-chain fatty acids, trimethylamine-N-oxide, and secondary bile acids^47-49^. Forth, influence on glucagon-like peptide-1 receptor in gut may generate impacts on blood pressure by regulation of atrial natriuretic peptide levels.^2^

Strengths of this study include large sample size, a relatively long follow-up of participants, adjustment for many important covariates (e.g., inflammatory indexes and use of proton pump inhibitors), and a comprehensive assessment of the associations between a wide range of gastroenterological disease and CVD risk and their interactions with age, sex, and BMI. Limitations deserve to be discussed when interpreting our findings. First, this observational study cannot infer causality of the identified associations between gastrointestinal diseases and the risk of incident CVD due to potential residual confounding (e.g. treatments without data) even though some of the associations have been verified in MR studies.^25,26,34^ Second, although the associations were robust in the analysis excluding incident CVDs within 2-year follow, our findings may still be influenced by reverse causality due to asymptomatic CVD. Third, there may be misclassifications of both the exposures and outcomes. However, this bias should be nondifferential due to the prospective design and thus attenuate the associations in a conservative way. Fourth, our findings were based on a British population of middled-aged and older participants. Whether these results can be generalized to other populations with largely different features needs confirmation.

In summary, this large-scale prospective cohort study identified associations of a broad range of gastrointestinal diseases with an increased risk of incident CVD, in particular CHD and PAD. The associations appeared to be stronger in women, younger individuals, and overweight and obese participants. Even though the associations were generally moderate except for that for liver disease, all these findings lend evidence support for the reinforced secondary prevention for CVD among patients with gastrointestinal disorders given a high prevalence of gastrointestinal disorders worldwide. More attention should be paid to young and overweight women with an excessive risk of CVD by the exposure to gastrointestinal diseases.

## Data Availability

The datasets analysed during the current study are available in a public, open-access repository

https://www.ukbiobank.ac.uk/

## Acknowledgments

We are much obliged to the administer team of the UK Biobank as well as all the participants.

## Data availability statement

The datasets analysed during the current study are available in a public, open-access repository (https://www.ukbiobank.ac.uk/).

## Ethics approval and consent to participate

The ethical approval was granted for the UK Biobank by the North West-Haydock Research Ethics Committee (REC reference: 21/NW/0157). All participants provided informed consent through electronic signature at baseline assessment. This study was conducted with the UK Biobank Resource under application number 66354.

## CRediT authorship contribution statement

All authors read and approved the final manuscript.

Jie Chen (Conceptualization: Equal; Methodology: Equal; Formal analysis: Supporting; Writing - original draft: Supporting; Writing - review & editing: Equal)

Yuhao Sun (Conceptualization: Equal; Methodology: Equal; Formal analysis: Leading; Writing - original draft: Supporting; Writing - review & editing: Equal)

Tian Fu (Conceptualization: Supporting; Methodology: Equal; Formal analysis: Supporting; Writing - original draft: Equal; Writing - review & editing: Equal)

Shiyuan Lu (Formal analysis: Supporting; Writing - original draft: Supporting; Writing - review & editing: Supporting)

Wenming Shi (Formal analysis: Supporting; Writing - original draft: Supporting; Writing - review & editing: Supporting)

Jianhui Zhao (Formal analysis: Supporting; Writing - original draft: Supporting; Writing - review & editing: Supporting)

Sen Li (Formal analysis: Supporting; Writing - original draft: Supporting; Writing - review & editing: Supporting)

Xue Li (Conceptualization: Equal; Data curation: Equal; and Funding acquisition: Leading; Writing - review & editing: Equal)

Shuai Yuan (Conceptualization: Leading; Methodology: Equal; Writing - review & editing: Leading)

Susanna C. Larsson (Conceptualization: Equal; Data curation: Equal; and Funding acquisition: Equal; and Writing - review & editing: Leading)

